# Trend, multivariate decomposition and spatial distribution of perinatal mortality in Ethiopia using further analysis of EDHS 2005-2016

**DOI:** 10.1101/2023.07.25.23293164

**Authors:** Muluken Chanie Agimas, Demewoz Kefale, Tigabu Kidie Tesfie, Demewoz Kefale, Worku Necho, Tigabu Munye, Gedefaw Abeje, Yohannes Tesfahun, Amare Simegn, Amare kassaw, Shegaw Zeleke, Solomon Demis, Habtamu Shimels Hailemeskel

## Abstract

**Background:** Perinatal mortality is the global health problem, especially Ethiopia has the highest perinatal mortality rate. Studies about perinatal mortality were conducted in Ethiopia, but which factors specifically contribute to the change in perinatal mortality across time is unknown.

**Objectives:** To assess the trend, multivariate decomposition and spatial analysis of perinatal mortality in Ethiopia using EDHS 2005 to 2016.

**Methods:** A community-based cross-sectional study design was used. EDHS 2005-2016 data was used and weighting has been applied to adjust the difference in the probability of selection. Logit based multivariate decomposition analysis was used using STATA version 14.1. Moran’s I statistics using ArcGIS was also used to identify the significant clustering of perinatal mortality.

**Result:** The trend of perinatal mortality in Ethiopia decreased from 37 per 1000 births in 2005 to 33 per 1000 births in 2016. About 83.3% of the decrease in perinatal mortality in the survey was attributed to the difference in the endowment (composition) of the women. Among the differences in the endowment, the difference in the composition of ANC visits, take TT vaccine, urban residence, occupation, secondary education, birth attendant significantly decreased the perinatal mortality in the last 10 years. Among the differences in coefficients, skilled birth attendant significantly decreased the perinatal mortality. The spatial distribution of perinatal mortality was randomly distributed.

**Conclusion:** The perinatal mortality in Ethiopia has declined over time. Variables like ANC visit, taking TT vaccine, urban residence, have occupation, secondary education and skilled birth attendant reduce perinatal mortality. Perinatal mortality was distributed randomly in Ethiopia. To reduce perinatal mortality more, scaling-up the maternal and newborn health services has a critical role.

## Introduction

Perinatal mortality is the death of a fetus starting at 28 weeks of gestation and newborn death up to 7 days after delivery (1, 2). The well-being of women and newborns can be measured by perinatal mortality (3). Goals are set by the world health organization and sustainable development goals to reduce the death of newborns by 2030 to less than 12 per 1000 live births (4, 5). By 2020, Ethiopia set up a plan to reduce perinatal mortality (PM) using improving maternal and newborn health services, such as improving antenatal care utilization, iron supplementation, and skilled delivery (6). Even though a lot of work has been done, the PM is still a major public health issue and a devastating problem.

About 6.3 million global PM are reported each year (3) and from this figure, 98% of the PM reported was from developing countries, especially in sub-Saharan African countries. PM is the most challenging problem (7). The geographic distribution of PM in Africa was in East Africa 58 per 1000 births, in middle Africa 75 per 1000 births, in Northern Africa 34 per 1000 births, in Southern Africa 37 per 1000 births, and in western Africa 76 per 1000 births (3). Among the sub-Saharan African countries, Ethiopia has the highest PM ranges from 66 to 124 per 1000 and 37 to 52 per 1000 total births in the health community-based setting respectively (11).Among the most critical times of maternal and newborn death, the first hour and week of delivery contribute to the highest death rate (8). To measure the reliable figure of mortality during delivery, PM is a more comprehensive indicator than the neonatal mortality indicator (9, 10).

The PM has several consequences. For example, psychological problems (depression, suicidal behavior) reduce work productivity because of grieving and negatively affect couples’ relationships (11, 12). Other mental disorders like anxiety/stress and poor partnership are also associated with perinatal loss (13). Quality of care during birth can save about three million still births and newborns each year (4).

Although interventions are implemented in Ethiopia to reverse the impact of the PM, the PM is still the most challenging public health problem (14). As different evidence shows; age, delivery place, sex of the child, residence of the women, antenatal care utilization status and source of drinking water were the significant factors for PM (15–18). Even though the PM is a critical problem in Ethiopia, the spatial distribution of the problem, the trend of PM using national level data (Ethiopian demographic health survey data), and the factors for the trend of PM were not addressed using decomposition analysis. Additionally, most studies about PM were conducted at health institution level but there are no adequate studies at the community level about the trend and spatial distribution of PM in Ethiopia. Therefore, the current study aimed to assess the spatial distribution, trend and determinants of the PM in Ethiopia using further analysis of Ethiopian demographic health survey (EDHS) 2005-2016 data set using decomposition and spatial analysis.

## Objectives

To determine the spatial distribution of perinatal mortality in Ethiopia using 2005 and 2011 EDHS data

To determine the trend of perinatal mortality in Ethiopia using EDHS 2005-2016 data.

To identify the factors attributed to the change in perinatal mortality in Ethiopia using decomposition analysis

## Methods

### Data

The EDHS data set of 2005, 2011 and 2016 were used for the intended analysis using community based Crossectional study design. These demographic surveys used stratified two stage cluster sampling techniques. For EDHS-2005, the sampling frame was developed based on the 1994 Ethiopian population and housing census. For EDHS-2011 and EDHS-201, the 2007 Ethiopian population and housing census was used for sampling frame preparation. Geographical areas or urban and rural areas were used for stratification purposes. About 21 sampling strata were used for the survey. A total of 540, 624 and 645 enumeration areas with independent and probability proportional for the enumeration areas were selected for EDHS-2005, 2011 and 2016 respectively. Then house hold listing and selection was conducted before the actual data collection procedure and, on average, 28 households in the enumeration areas were selected using systematic sampling. The detailed selection process was reported in EDHS-2005, EDHS-2011 and EDHS-2016 (19–21). A total of 33,998 sample size was used in all surveys (EDHS-2005=12035, EDHS-2011=10588, EDHS-2016=11375).

### Study and source population

The study population was all births from reproductive-age women in Ethiopia. Whereas, the study population was the selected enumeration areas in Ethiopia.

### Outcome of interest

Perinatal mortality (Yes, No)

### Operational definition

Perinatal mortality is the death of a fetus starting at 28 weeks of gestation and newborn death up to 7 days after delivery (1, 2).

### Data collection procedure

The EDHS data set was used for the current study. This data was accessed by requesting the DHS program website www.measuredhs.com and by explaining the ultimate objective of the current study. The raw data was collected from reproductive age women who gave birth using a pretested structured questionnaire. The data was extracted from birth records of the three consecutive EDHS data sets (2005–2016).

### Data management analysis

Before descriptive and analytical analysis, data were weighted using sampling weight techniques to be more representative. To describe the study subjects/ participants, descriptive measures like cross tabulation was used using STATA version-14. After important predictors are cleaned, coded and extracted, then EDHS 2005, 2011, 2016 data sets were merged together for further spatial trend and decomposition analysis. The Moran I statistics were analyzed using ArcGIS software version 10.3. The spatial distribution of perinatal mortality was analyzed for EDHS 2005 and 2011. Because spatial distribution for EDHS 2016 was already conducted from the previous study (22).

### Trend and decomposition analysis

To assess the trend of perinatal mortality, descriptive analysis was used using selected variables and a combination of EDHS data separately (2005–2011, 2011–2016, and 2005–2016). To determine the trend of perinatal mortality changes and the possible contributing predictors and the source of difference in perinatal mortality percentage in Ethiopia from 2005-2016, multivariate decomposition analysis was employed. Two important components, such as the composition of the population and the coefficient, were considered to be the contributing factors for the decreasing or increasing pattern of perinatal mortality in Ethiopia since 2005 to 2016. The logit based multivariate decomposition analysis was used to identify the possible factors for the trend of perinatal mortality in Ethiopia. The Logit based decomposition was using the STATA command mvdcmp (23). The observed difference in perinatal mortality between the surveys decomposed in to endowment (E) and coefficient (C) components. The endowment (E) means the difference in characteristics, whereas the coefficient (C) means the difference in effects.

### Spatial autocorrelation

The ArcGIS v-10.3 software was used to estimate Morn’s I spatial autocorrelation perinatal mortality in Ethiopia using EDHS 2005 and 2011 because spatial distribution using EDHS 2016 was conducted. The value of Moran’s I ranges from −1 to +1 and the value of −1 infers the dispersed variation of perinatal mortality. Whereas, +1 indicates that the clustered perinatal mortality and 0 Morn’s I test value showed the random distribution of perinatal mortality. To declare the statistical significance of the Moran’s spatial autocorrelation, a p-value of less than 0.05 was considered. Finally, the spatial autocorrelation was not statistically significant and the assumption of further spatial analysis was violated.

## Results

### Socio-demographic characteristics of the participants

In all surveys, 16853 (49.6%) of the participants were aged 20-29 years. Regarding employment status, there was a slight decrement in no work (unemployment) from 2177 (18.1%) in 2005 EDHS, to 1457 (12.8%) in 2016 EDHS. From all surveys, about 18439 (54.2%) participants have had an occupation. Additionally, relatively the highest 4997 (43.9%) of the participants were included in the 2016 EDHS. Furthermore, the majority of participants, 24551 (72.2%) those who were included in the three surveys, had no education and there was a decrement from 9541(79.3%) in 2005 to 7606 (66.9%) in 2016 **(Table-1).**

**Table-1:** socio demographic characteristics of the participants in 2005, 2011 and 2016 EDHS.

| Variable |  | Weighted frequency (%) |  |  |
| --- | --- | --- | --- | --- |
|  |  | 2005(N=12037) | 2011(N=10588) | 2016(N=11375) |
| Women age | <20 | 576 (4.8%) | 361 (3.4%) | 374 (3.3%) |
|  | 20-29 | 5825 (48.4%) | 5429 (51.3%) | 5599 (49.2%) |
|  | 30-39 | 4344 (36.1%) | 3823 (36%) | 4382 (38.5%) |
|  | 40-49 | 1292 (10.7%) | 985 (9.3%) | 1020 (9%) |
| Residence | Urban | 883 (7.3%) | 1406 (13.3%) | 1229 (10.8%) |
|  | Rural | 11,154 (92.7%) | 9182 (86.7%) | 10,146 (89.2%) |
| Region | Tigray | 775 (6.4%) | 678 (6.4%) | 709 (6.2%) |
|  | Afar | 122 (0.01%) | 114 (1.1%) | 119 (1%) |
|  | Amhara | 2827(23.5%) | 2538 (24%) | 2121 (18.6%) |
|  | Oromia | 4666 (38.8%) | 4291 (40.6%) | 4997 (43.9%) |
|  | Somalia | 522 (4.3%) | 296 (2.8%) | 554 (4.9%) |
|  | Benishangul | 115 (0.9%) | 131 (1.2%) | 133(1.3%) |
|  | SNNPRs | 2742 (22.8%) | 2248 (21.2%) | 2402 (21.1%) |
|  | Harari | 23 (0.2%) | 27 (0.2%) | 27 (0.2%) |
|  | Addis Ababa | 170 (1.5%) | 192 (1.8%) | 234 (2.1%) |
|  | Dire Dawa | 41 (0.3%) | 35 (0.3%) | 49 (0.4%) |
|  | Gambela | 34 (0.03%) | 38 (0.4%) | 29 (0.3%) |
| Religion | Orthodox | 5094 (42.3%) | 4233 (40%) | 3844 (33.8%) |
|  | Muslim | 4123 (34.3%) | 3563 (33.7%) | 4696 (41.3%) |
|  | Others | 2820 (23.4%) | 2792 (26.3%) | 2835 (24.9%) |
| Maternal educational level | No education | 9541 (79.3%) | 7404 (69.9%) | 7606 (66.9%) |
|  | Primary education | 1993 (16.5%) | 2801(26.5%) | 2961 (26%) |
|  | Secondary and higher education | 503 (4.2%) | 383 (3.6%) | 808 (7.1%) |
| Wealth index | Poorest | 2646 (22%) | 2421(22.9%) | 2483 (21.8%) |
|  | Poorer | 2497(20.7%) | 2382 (22.5%) | 2877 (25.3%) |
|  | Middle | 2607 (21.7%) | 2151 (20.3%) | 2318 (20.4%) |
|  | Richer | 2468 (20.5%) | 2018(19.1%) | 2084 (18.3%) |
|  | Richest | 1819 (15.1%) | 1616 (15.3%) | 1613 (14.2%) |
| Parent's<br>education | No education | 3458(28.7%) | 2598 (24.5%) | 3201(28.1%) |
|  | Primary | 4568 (38%) | 6201(58.6%) | 4562(40.1%) |
|  | Secondary<br>education | 4011(33.3%) | 1789 (16.9%) | 3612 (31.8%) |
| Parent's<br>occupatio<br>n | No work | 2177(18.1%) | 2346 (22.2%) | 1457 (12.8%) |
|  | Non agricultural | 6052(50.3%) | 5468 (51.6%) | 5901(51.9%) |
|  | Agricultural | 3808(31.6%) | 2774 (26.2%) | 4017 (35.3%) |
| Mother<br>occupatio<br>n | No occupation | 4348 (36%) | 4861(45.9%) | 6352 (55.8%) |
|  | Had occupation | 7689 (64%) | 5727 (54.1%) | 5023 (44.2%) |

### The trend of perinatal mortality rate in Ethiopia

The rate of perinatal mortality in Ethiopia has significantly decreased from a 37 death rate per 1000 births in 2005 to a 33 death rate per 1000 births in 2016 **(Figure-1).**

**Figure-1:**
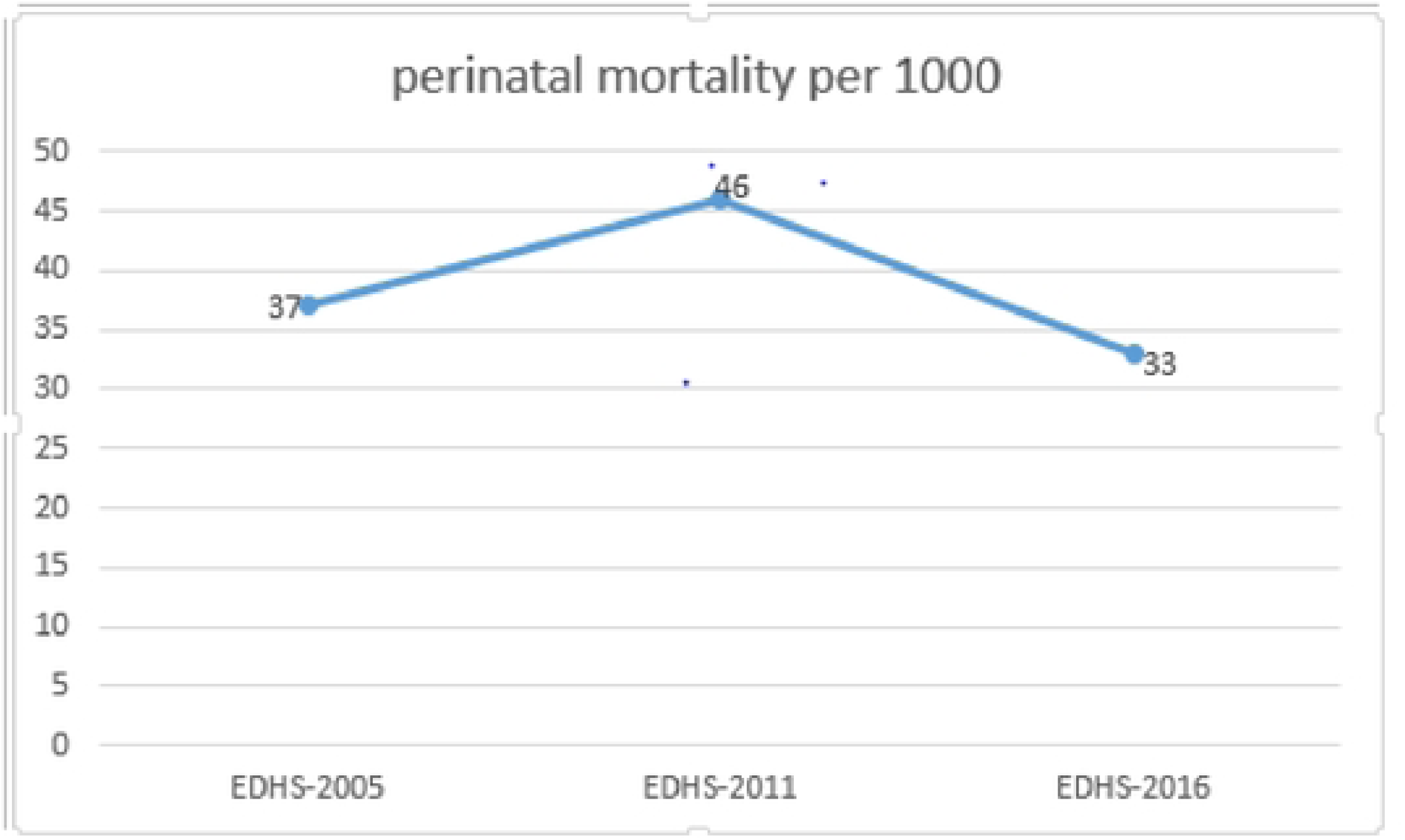
trend of perinatal mortality in Ethiopia from EDHS 2005 to EDHS 2016.

### Environmental and health service related characteristics

About 11154 (92.7%), 9182 (86.7%) and 10750 (94.5%) of the participants used toilet facilities in 2005, 2011 and 2016 respectively, and there was an increment in toilet facility utilization from 11154 (92.7%) in 2005 to 10750 (94.5%) in 2016. Skilled births were increased from 5147 (51%) in 2005 to 6563 (62%) in 2011 to 8697 (76.5%) in 2016. The overall ANC utilization in all surveys was 20635 (61%) and the magnitude was increased from 43.3% in 2005 to 61.6% in 2011 to 78.2% in 2016 (**Table-2).**

**Table-2:** distribution of environmental and health service related characteristics of respondents in 2005, 2011 and 2016 EDHS.

| Variable |  | Weighted frequency (%) |  |  |
| --- | --- | --- | --- | --- |
|  |  | 2005(N=12037) | 2011(N=10588) | 2016(N=11375) |
| Source of drinking water | Piped | 2576(21.4%) | 1361 (12.9%) | 1924 (16.9%) |
|  | Protected | 5825 (48.4%) | 5404 (51%) | 5519 (48.5%) |
|  | Unprotected | 3636 (30.2%) | 3823 (36.1%) | 3932 (34.6%) |
| Use of toilet facilities | Yes | 11154 (92.7%) | 9182 (86.7%) | 10750 (94.5%) |
|  | No | 883 (7.3%) | 1406 (13.3%) | 625 (5.5%) |
| Type of cooking fuel | Solid | 7752 (64.4%) | 6789 (64%) | 6701 (58.9%) |
|  | Non solid | 4285(35.6%) | 3789 (36%) | 4674 (41.1%) |
| Delivery assistance | Non skilled | 5890 (49%) | 4025 (38%) | 2678 (23.5%) |
|  | Skilled | 5147 (51%) | 6563 (62%) | 8697 (76.5%) |
| Mode of delivery | Vaginal | 10036 (83.4%) | 7687 (72.6%) | 8606 (75.7%) |
|  | Cesarean-section | 2001 (16.6%) | 2901 (27.4%) | 2769 (24.3%) |
| ANC utilization | Yes | 5211(43.3%) | 6524 (61.6%) | 8900 (78.2%) |
|  | No | 6826 (56.7%) | 4064 (38.4%) | 2475 (21.8%) |
| Tetanus vaccination status | Not use | 2123(17.7%) | 2620 (24.7%) | 1890 (16.6%) |
|  | 1 dose | 5579(45.3%) | 4900 (46.3%) | 4005 (35.2%) |
|  | ≥2 doses | 4335(36%) | 3068 (29%) | 4480 (39.4%) |

### Fetal and neonatal related characteristics

The majority of the births, 10836 (90%) in 2005, 9602 (90.7%) in 2011 and 10186 (89.5%) in 2016 were single. More than half of 23447 (69%) the births interval in all survey was 24 months and above. Furthermore, there was a decrement in cigarette smoking from 256 (21.3%) in 2005 to 189 (17.8%) in 2011 and 126 (1.1%) in 2016 **(Table-3).**

**Table-3:** Fetal/neonatal and maternal behavior related characteristics of respondents in 2005, 2011 and 2016 EDHS.

| Variable |  | Weighted frequency (%) |  |  |
| --- | --- | --- | --- | --- |
|  |  | 2005(N=12037) | 2011(N=10588) | 2016(N=11375) |
| Birth interval | <24 months | 4212 (35%) | 3565 (33.7%) | 2776 (24.4%) |
|  | ≥24 months | 7825 (65%) | 7023 (66.3%) | 8599 (75.6%) |
| Sex of child | Female | 5987 (49.7%) | 5067 (47.9%) | 5897 (51.8%) |
|  | Male | 6050 (50.3%) | 5521(52.1%) | 5478 (49.2%) |
| Birth order | ≤3 | 3679 (30.6%) | 3101((29.3%) | 2989 (26.3%) |
|  | 4-5 | 4356 (36.2%) | 3780 (35.7%) | 5231(46%) |
|  | ≥6 | 4002 (33.2%) | 3707 (35%) | 3155 (27.7%) |
| Types of births | Single | 10836 (90%) | 9602 (90.7%) | 10186 (89.5%) |
|  | Multiple | 1201 (10%) | 986 (9.3%) | 1189 (10.5%) |
| Smoking cigarette | Yes | 256 (21.3%) | 189 (17.8%) | 126 (1.1%) |
|  | No | 11781(78.7%) | 10399 (81.2%) | 11249 (98.9%) |

### Trends of perinatal mortality rate by women characteristics

The trend of perinatal mortality rate by women’s characteristics in Ethiopia from 2005-2016 EDHS showed that there was a decline in perinatal mortality in urban residences across all surveys from 23.3 per 1000 births in 2005 to 16.6 per 1000 births in 2011 and 6.6 per 1000 in 2016. Regarding the regional distribution of perinatal mortality rate, in the Amhara region, the trend decline across all phases in 2005-2011 (phase-I) declined by 29.9 per 1000 births. In 2011-2016 (phase-II), declined by 10.6 per 1000 births and the maximum amount of change was observed in phase III (2005–2016) which was declined by a death rate of 40.5 per 1000 births. Furthermore, the trend of perinatal mortality rate by women aged less than 20 years showed that in phase I (2011–2005) it was decreased by 1.6 per 1000 births, in phase III (2016–2011) by 3.4 per 1000 births and by 5 per 1000 births in phase III (2016–2005) **(Table-4).**

**Table-4:**
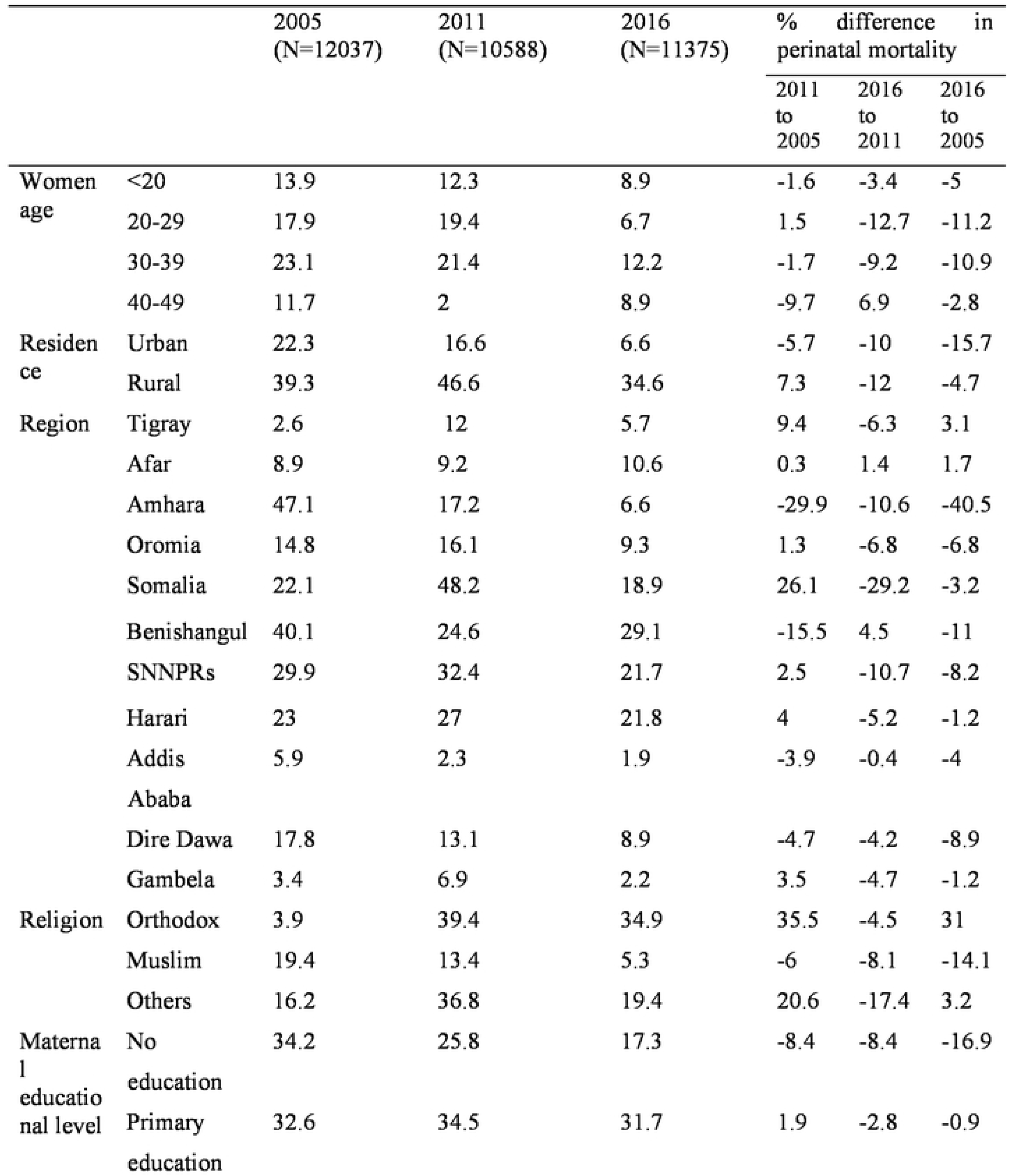
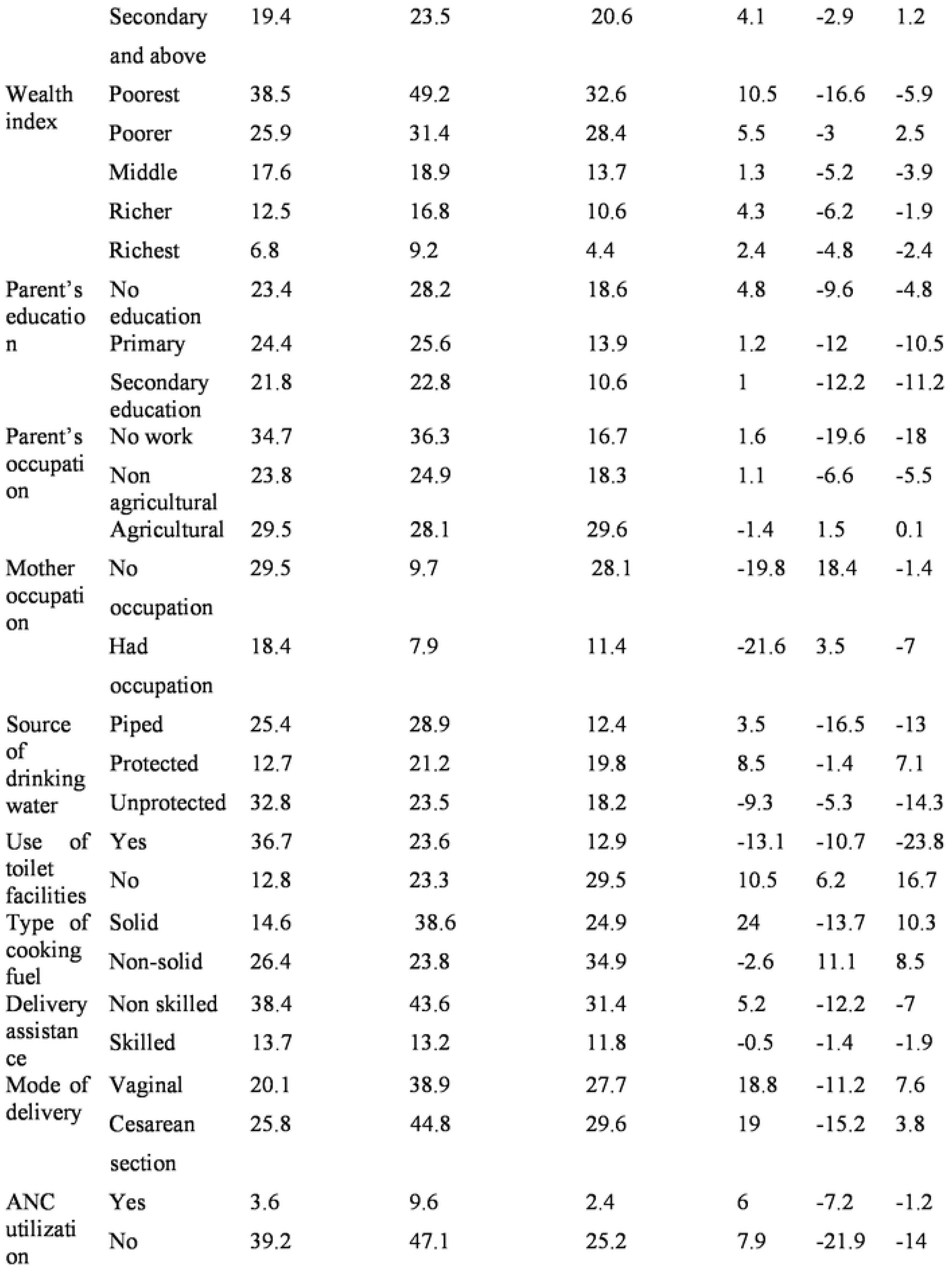
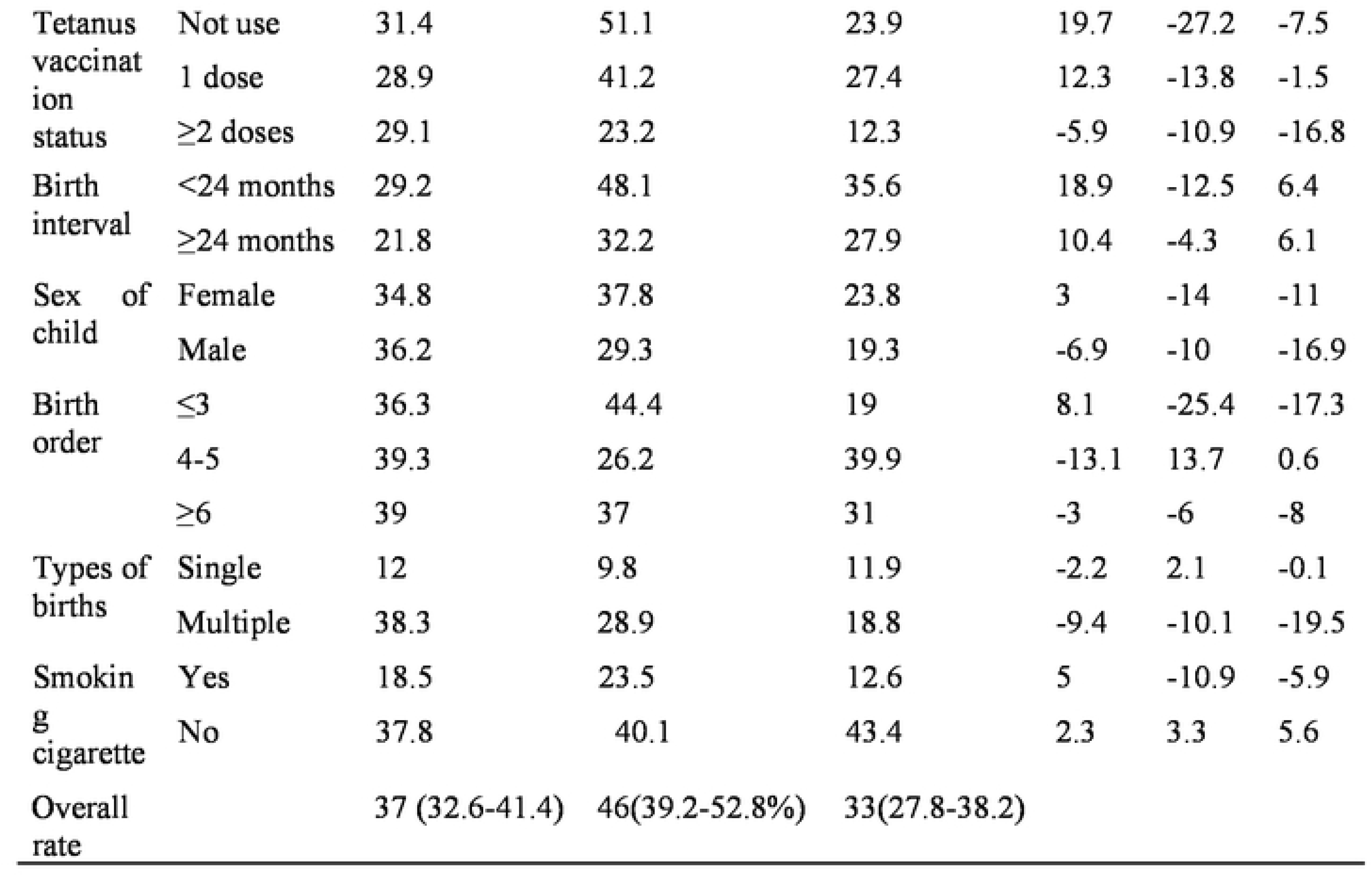
Trends in perinatal mortality rate among women’s who gave birth in the last live years prior to the surveys by selected characteristics 2005, 2011, and 2016 EDHS.

### Spatial distribution of perinatal mortality in Ethiopia

Before conducting spatial interpolation, Sat scan analysis, hot/cold spot analysis and other spatial analysis, the spatial autocorrelation analysis (global Moran’s I test) was conducted on EDHS 2005 and 2011. Thus, the global Moran’s I test, evidenced that the spatial distribution of perinatal mortality in Ethiopia was randomly distributed. The Moran’s I, z-value and p-value of perinatal mortality distribution (for EDHS-2005: Moran’s I = −0.05771, Z-value= −0.269571, p-value= 0.787490, for EDHS-2011: Moran’s I = −0.027777, Z-value= −0.473613, p-value= 0.635776) **(fig-2 and fig-3).** This finding implies that perinatal mortality in Ethiopia was not clustered or the distribution was just random. Therefore, further spatial analysis was impossible to conduct.

**Fig-2:**
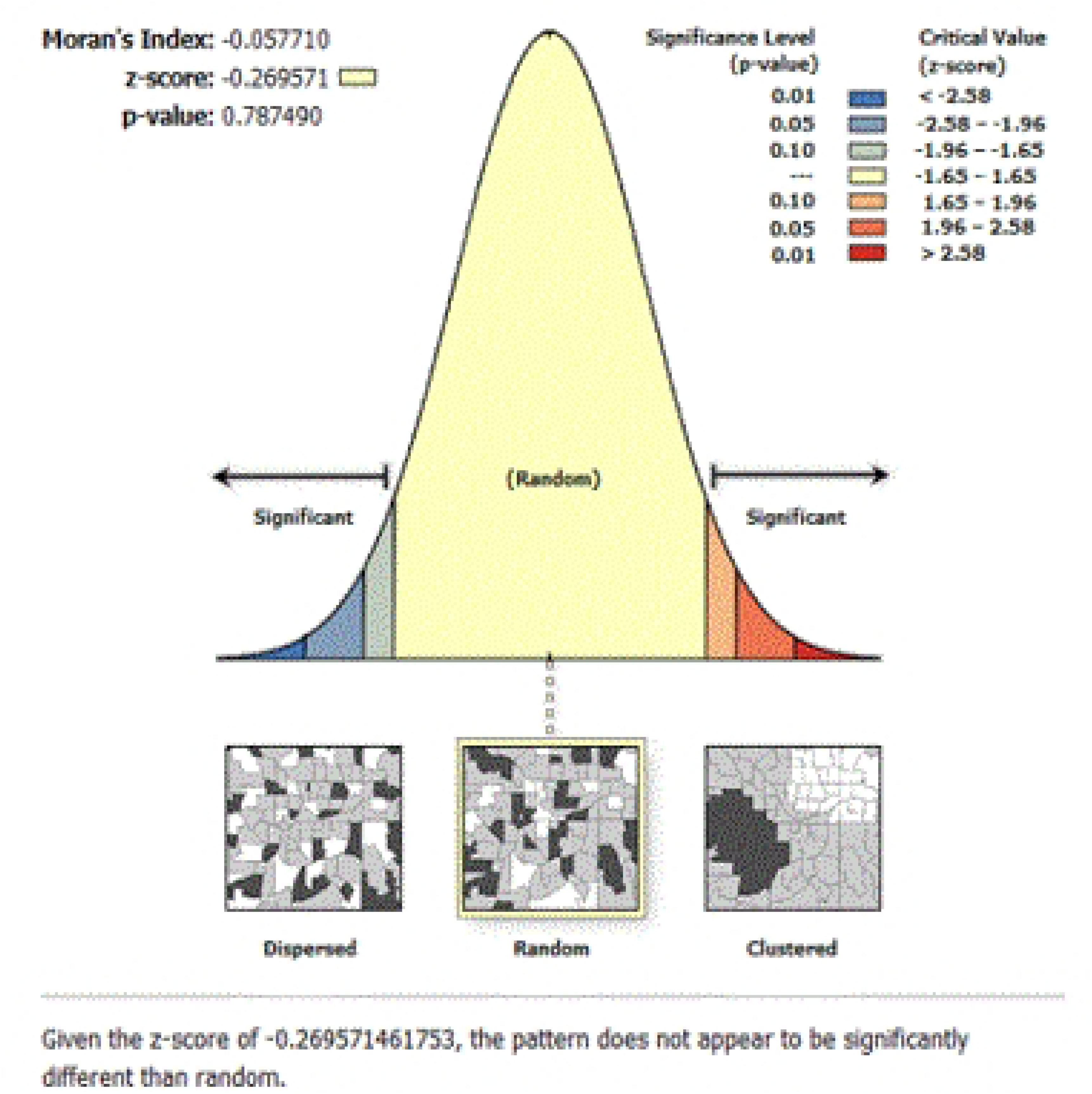
Spatial autocorrelation of perinatal mortality in Ethiopia using 2005 EDHS.

**Fig-3:**
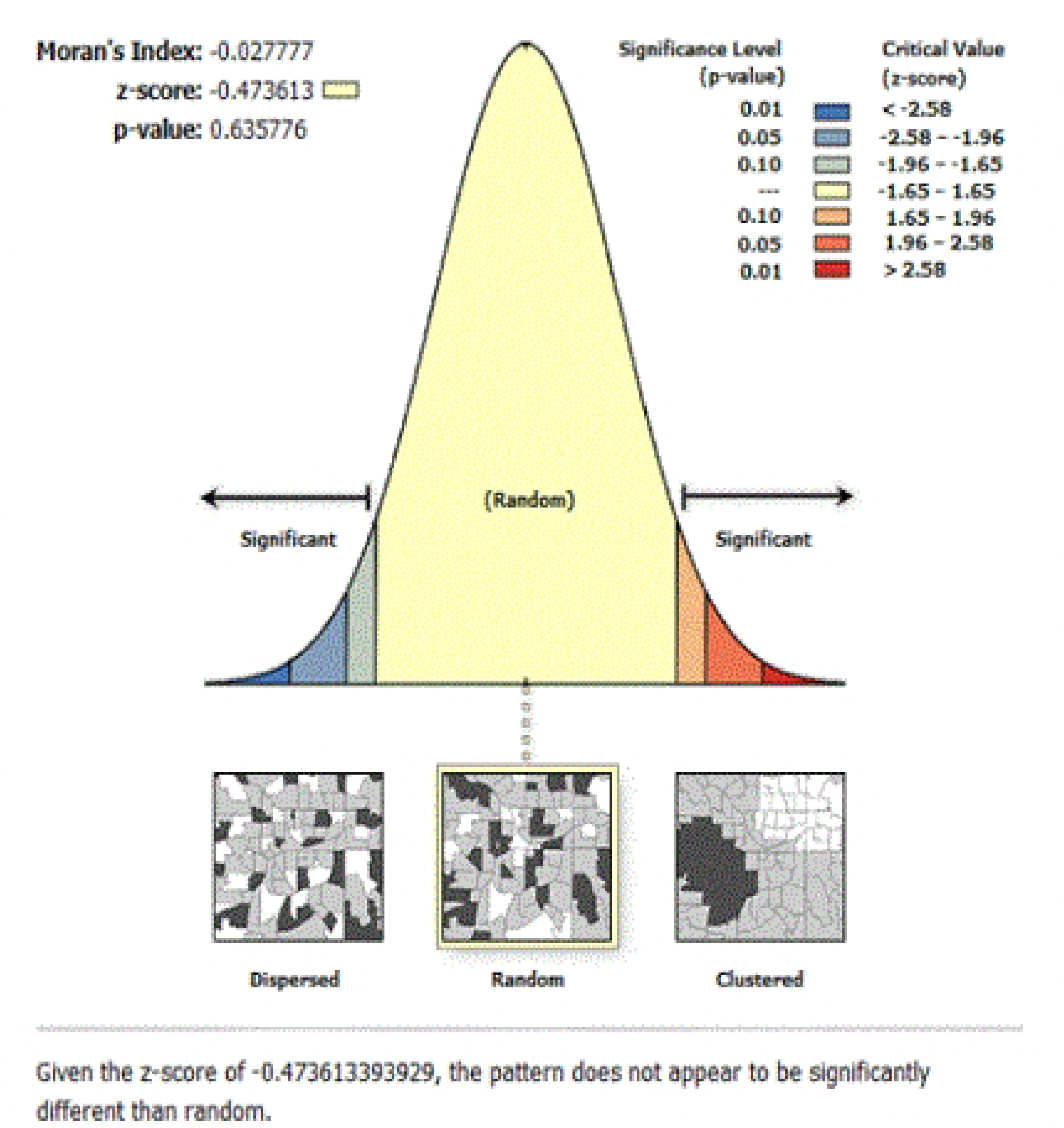
Spatial autocorrelation of perinatal mortality in Ethiopia using 2011 EDHS.

### Over all decomposition analysis

The overall perinatal mortality in Ethiopia has significantly decreased from 2005 to 2016. As the overall decomposition (2005–2016) analysis showed, the decrement of perinatal mortality in Ethiopia was explained by the endowment of the women between each EDHS. Thus, around 83.3 % of the perinatal mortality decline was because of the endowment (composition) of the participants and which was statistically significant. The rest was attributed to the change in the coefficients (C) **(Table-5).**

**Table-5:** Overall decomposition analysis of change in perinatal mortality in Ethiopia 2005-2016.

| Perinatal mortality | Coefficient | 95% CI | p-value | Pct. |
| --- | --- | --- | --- | --- |
| **E** | -0.0083 | (-0.0157, -0.001) | 0.026 | 83.3 |
| **C** | -0.00167 | (-0.0031,-0.00025) | 0.02 | 16.7 |
| **R** | -0.01 | -0.017, -0.0029 | 0.006 |  |
| Note E= Endowment<br>C=Coefficient<br>R=Residual |  |  |  |  |

### Detailed decomposition analysis

Among the difference in the endowment; the difference in the composition of ANC visit (*B*= −0.002164, 95%CI; −0.0035, −0.00358, p-value=0.001), take one TT dose (*B*= −0.00072, 95% CI; −0.0012, −0.0011, p-value=0.007), ≥2 TT vaccine (*B*=-0.0019 95% CI; −0.0028, −0.0011, p-value=<0.001), Urban residence (*B*=-0.000499, 95% CI; −0.0009, −0.00007, p-value=0.02), have occupation (*B*=-0.019, 95%CI; −0.0033, −0.00061, p-value= 0.004), secondary education (B= 0.0023, 95% CI; 0.000046, 0.0041, p-value=0,014), skill birth attendant (*B*=0.0069, 95% CI; 0.00068, 0.013, p-value= 0.03) were significantly decrease the perinatal mortality in the last 10 years. Among the differences in coefficients, skilled birth attendant (*B*=0.000099, 95% CI; 0.00002, 0.00018, p-value=0.013) significantly decreased the perinatal mortality **(Table-6).**

**Table-6:** detailed decomposition analysis of perinatal mortality in Ethiopia from 2005−2016.

| Experiencing perinatal mortality |  | Difference due to characteristics (E) |  |  | Difference due to coefficient (C) |  |  |
| --- | --- | --- | --- | --- | --- | --- | --- |
|  |  | Coef. | p-value | Pct. | Coef. | p-value | Pct. |
| Mode of delivery | Vaginal delivery |  |  |  |  |  |  |
|  | Cesarean section | -0.000042<br>(-0.00039, 0.00031) | 0.59 | 2.8 | -0.00031<br>(-0.0008,0.0014) | 0.812 | -0.39 |
| ANC | Yes | -0.002164<br>(-0.0035, -0.00358) | 0.001 | -19.83 | -0.00358<br>(-0.0089,0.0018) | 0.193 | -32.84 |
|  | No |  |  |  |  |  |  |
| TT vaccine uptake status | 1 | -0.00072<br>(-0.0012, -0.0011) | 0.007 | -6.6 | -0.00036<br>(-0.0028,0.0021) | 0.77 | -3.38 |
|  | ≥2 | -0.0019<br>(-0.0028, -0.0011) | <0.001 | -17.8 | -0.0011<br>(-0.0064,-0041) | 0.67 | -10.44 |
|  | No |  |  |  |  |  |  |
| Place of residence | Urban | -0.000499<br>(-0.0009, -0.00007) | 0.02 | -4.59 | -0.0073<br>(-0.01, 0.025) | 0.419 | 65.24 |
|  | Rural |  |  |  |  |  |  |
| Maternal occupation | Occupation | -0.019<br>(-0.0033, -0.00061) | 0.004 | -18.1 | 0.0082<br>(-0.0043,0.021) | 0.2 | 75.13 |
|  | No occupation |  |  |  |  |  |  |
| Maternal education | No education |  |  |  |  |  |  |
|  | Primary education | 0.00033<br>(-0.0002, 0.0027) | 0.78 | -0.33 | -0.00094<br>(-0.0041,0.0022) | 0.56 | 9.4 |
|  | Secondary education | 0.0023<br>(0.000046, 0.0041) | 0.014 | -2.31 | -0.0016<br>(-0.0074,0.0041) | 0.58 | 16.4 |
| Delivery assistance | Skilled | 0.0069<br>(0.00068, 0.013) | 0.03 | -69 | 0.000099<br>(0.00002,0.00018) | 0.013 | -0.98 |
|  | Non skilled |  |  |  |  |  |  |
| Total |  |  |  | 83.3 |  |  | 16.7 |
| Constant |  |  |  |  | -0.0045<br>(-0.0023,-0.0098) |  |  |

## Discussion

Perinatal mortality is the most devastating issue in Ethiopia. But the trend, spatial distribution and what factors contributed to the increment or decrement of perinatal mortality were not studied. Thus, in this study, an attempt has been made to assess the trend, multivariate decomposition and spatial distribution of perinatal mortality in Ethiopia.

The trend of perinatal mortality in Ethiopia has decreased over time from 37 deaths per 1000 births in 2005 to 34 deaths per 1000 births in 2016. This was supported by a systematic review and meta-analysis conducted in Ethiopia (24). This may be because the Ethiopian government gave an emphasis on maternal and child health services by strengthening the health extension program and by exempting the service (25). The other possible reason might be associated with the numbers of skilled birth attendants and the basic service coverage like ANC, post-natal care service, family planning service, and institutional delivery service in Ethiopia increased over time, which enabled the women to get counseling about their pregnancy, their health, fetal condition, birth preparedness, nutrition and other important messages (26, 27).

As Moran’s I statistics test value showed, the spatial distribution of perinatal mortality in Ethiopia was not clustered or the pattern of perinatal mortality in Ethiopia was a result of random chance. Regarding the multivariate decomposition findings, the overall contribution of women’s composition for perinatal mortality in the entire survey (2005–2016) was 83.3%. As an increase in the endowment of urban resident women over the survey, there was a significant decrease in perinatal mortality in Ethiopia. This may be because the expansion of urbanization in Ethiopia may increase the chance of getting maternal and child health services and access to health information more than rural women (28, 29).

The increase in the women’s composition of ANC utilization over the survey also had a significant effect on the decrement in perinatal mortality in Ethiopia. This may be because women who visit for ANC service are more likely to give live birth and children have more survival than women who have no history of ANC service (30). An increase in taking the TT vaccine (taking one or more TT vaccines) over the survey decreased the perinatal mortality significantly. The possible reason that might be associated with improving the TT vaccination coverage could be to substantially increase the number of newborns protected at birth than their counterparts and subsequently reduce the number of newborn deaths (31). The other possible reason might be because those women who take the TT vaccine are more likely to access other health services like education about pregnancy complications, danger signs, nutrition counselling and other key messages.

An increase in women who had occupations also had a significant effect on the reduction of perinatal mortality in Ethiopia. This might be because women who have an occupation are more likely to become employed and live in a better socio-economic condition and this leads to better household food security and may reduce maternal malnutrition and its complications (32). The other possible reason might be that women who have an occupation are more likely to have better knowledge and educational status than those who have no occupation. Thus, better knowledge and educational background is a key to having better information about their health and having good health-seeking behavior for maternal and child health services.

In the current study, an increase in endowment of women with secondary education also significantly reduced the perinatal mortality compared to women with no education. This might be due to women with better educational status, which may increase the chance of having better health information, better nutrition, and better health-seeking behavior than those women who have no education in return, which improves infant survival and healthy maternal condition (33). Furthermore, an increase in the endowment (composition) of women who give birth with the assistance of a skilled birth attendant significantly reduces the perinatal mortality compared to women who give birth to a non-skilled birth attendant. Regarding the difference of coefficient, giving birth by a skilled birth attendant also significantly contributed to the reduction of perinatal mortality in Ethiopia. The finding was consistent with a systematic and meta-analysis study and a study conducted in the United States of America (34, 35). This can be justified by women who give birth by skilled birth attendant are get better delivery care than the counterparts. This can be justified by women who give birth by skilled birth attendants are get better delivery care than the counterparts. This could reduce the chance of newborn death by sepsis, hypothermia, prolonged labor and other complications during giving birth. The limitation of the study was some of the most important variables such as clinical data, maternal medical and obstetric condition and RH incompatibility, were missed in the EDHS data. So further study by considering the above factors is very important.

## Conclusion

The trend of perinatal mortality in Ethiopia has declined across time. The change in women’s endowment contributes to more than a third quarter of the perinatal mortality decrease. Among the change in women’s endowment, variables such as the uptake of the TT vaccine, skilled birth attendant, secondary education, occupation and ANC utilization, contributed to the decline of PM. From the change in difference coefficients, give birth by a skilled birth attendant was the significant predictor for the decline of PM in Ethiopia. To further reduce the PM, scaling-up the maternal health service has an abundant advantage. The finding has an implication for designing PM reduction strategies and wise allocation of resources for the interventions.

## Ethical declaration

### Ethical approval and consent to participate

Ethical approval from individual participants was not obtained because of secondary data. Rather permission was obtained from DHS international to access the data.

### Consent for publication

it is not applicable

### Data availability statement

All data are accessed in the article

## Data Availability

All relevant data are within the manuscript and its Supporting Information files.

## Acknowledgments

The authors would like to give thanks to DHS international for giving permission to utilize the data.

## Contribution of the Authors

**Conceptualization**: Muluken Chanie, Solomon Demis, Worku Necho, Habtamu Shimels Hailemeskel

**Data curation**: Solomon Demis, Amare Simegn, Tigabu Munye, Yohannes Tesfahun

**Formal analysis:** Muluken Chanie Agimas, Amare Kassaw, Amare Simegn, Demewoz Kefale

**Investigation**: Muluken Chanie Agimas, Tigabu Kidie, Tigabu Munye, Yohannes Tesfahun

**Methodology**: Muluken Chanie Agimas, Demewoz Kefale, Worku Necho, Gedefaw Abeje

**Software**: Muluken Chanie Agimas, Yohannes Tesfahun, Amare Simegn, Shegaw Zeleke, Habtamu Shimels Hailemeskel

**Supervision**: Muluken Chanie Agimas, Habtamu Shimels Hailemeskel

**Visualization**: Shegaw Zeleke, Solomon Demis, Tigabu Munye, Tigabu Kidie, Yohanes Tesfahun, Worku Necho, Demewoz Kefale, Gedefaw Abeje

**Writing – original draft**: Muluken Chanie Agimas, Solomon Demis, Shegaw Zeleke, Gedefaw Abeje

## Abbreviation and acronym

ANC……Antenatal Care

DHS……Demographic Health Survey

EDHS…..Ethiopian Demographic Health Survey

PM…..Perinatal Mortality

TT……Tetanus Toxoid

